# Improving automated prostate pathological grading via confidence filtering

**DOI:** 10.1101/2025.11.25.25340482

**Authors:** Ryan B. Fogarty, Dmitry B. Goldgof, Lawrence O. Hall, Jasreman Dhillon, Vaibhav Chumbalkar, Yoganand Balagurunathan

**Affiliations:** Department of Machine Learning, Moffitt Cancer Center, Tampa, FL, USA; Bellini College of Artificial Intelligence, Cybersecurity and Computing, University of South Florida, Tampa, FL, USA; Department of Pathology, Moffitt Cancer Center, Tampa, FL, USA

## Abstract

There have been many promising developments in deep learning to identify degrees of malignancies in prostate cancer pathologies. Deep network models have been shown to be useful in identifying patterns in histology images assessed at different scales.

Prostate pathological grade identification has been a challenge among clinical experts due to complex patterns on the whole slide level, for hematoxylin and eosin (H&E) stained samples. In this study, we identify primary patterns (Gleason) in small sections of the whole slide composed of uniform glandular patterns. We then follow sample selection methods that eliminate ambiguous regions or tiled-samples by confidence filtering. A pseudo-confidence is derived from the predicted output of the network, which is used as a quality indicator to consider the sample for discriminatory analysis.

We provide further evidence that using highly calibrated confidence sample selection, these gland-level features on the prostate biopsy sections can discriminate degrees of malignancy following primary Gleason patterns.

We used an optimized deep network (convolutional neural network, CNN) discriminating glandular regions with aggressive grades (Gleason 3 from 4) showed an accuracy of 0.68(0.04), F_1_ of 0.66(0.06) and AUC of 0.74(0.04). We further improve this result using confidence filtering, with a sample fraction of 0.35 (with a calibrated confidence of greater than 0.85), achieving an accuracy of 0.74 (0.08), F_1_ of 0.72 (0.12), and AUC 0.79 (0.08) averaged from holdout sets over multiple reshuffled experiments.

## Introduction

Pathological assessment of prostate malignancy follows International Society for Urological Pathology (ISUP) grade patterns, which describe the extent of neoplasm on hematoxylin and eosin (H&E) slides [1]. The interpretation difficulty of complex histopathological specimens is evident, with a high level of variability among pathologists. It has been well documented in several studies that have shown lower concordance between the pathologists’ opinion (intra and inter-institutional) [2]. There is a need for standardized clinical evaluation to improve the grading and clinical care. Differences in interpretation of clinical grades in histopathology slides are due to both clinical opinion differences and data sampling, leading to aleatoric noise in data labels, which is a consequence of pattern complexity; methods that do not consider these label errors will affect the machine learning model performance.

In earlier work, deep learning methods have been proposed to assist in Gleason pattern grading of digital whole slide images (WSI) without addressing clinical label level noise; these methods continue to be well-investigated [3–7]. Most research has concentrated on evaluating entire specimens or identifying Gleason grades or scores over large areas of the digitized biopsy. Due to the high resolution of digitized WSIs (typically ranging from 20x-40x or 50-100 mega-pixels), vision algorithms require the image to be tiled into many smaller patches that are evaluated independently and then encoded into a much smaller latent space.

Behzadi et al. used a transformer and multi-instance learning (MIL) to classify pathology features. The derived patches were shown to be good predictors of pathological grade. State-of-the-art performance was achieved in classifying entire biopsies by selecting the most relevant patches of the WSI with the use of an autoencoder and trained attention network [4]. Generalization performance has improved by utilizing the diversity and amount of data available with TCGA [8], PANDA [9] and Gleason 2019 [10, 11] datasets.

Xu et al have created an extremely large pathology dataset with over 1.3 billion 256x256 patches derived from 117 thousand whole slide images collected from more than 30,000 patients at the Providence group in Portland OR, and Seattle WA [5].

A study by Butt et al. demonstrated that applying multiple Gleason scores to each patch or multi-labeling improves overall pathological grading by estimating pixel labeling error of up to 30% on the SICAPv2 pathology dataset [6].

The Deep Gleason study has demonstrated state-of-the-art performance with CNN architectures for Gleason scoring on whole slide images utilizing the ConvNeXt network, which has been shown to be even more effective than modern transformer architectures such as the Swin (sliding windows) transformer [7, 12].

Our prior study [13] demonstrated that the random sampling and consensus (RANSAC) technique [14] provides a framework to handle class mislabeling. We presented a deep model to discriminate primary Gleason patterns using well-curated patterns in representative regions (with patch areas of ∼ 0.14*mm*^2^), which reported an accuracy of 0.97 and *F*_1_-score of 0.97 (using Monte-Carlo Cross Validation). The RANSAC approach is comparable to “confident learning” [15], recently developed techniques that are available in the Cleanlab software library [16]. A generalized approach has recently been summarized in a label-correction framework [17].

Several approaches to confidence calibration have been developed to provide meaningful confidence intervals to inform the clinician and patient. Jiang et al. compare several conventional techniques such as histogram binning, Platt scaling, and isotonic regression to their own adaptive calibration of predictions (ACP) method [18]. Gawlikowski et al. provide a survey of techniques and discussion of the various forms of aleatoric and epistemic error and some approaches to minimize these errors [19]. Our study demonstrates the Parzen-Rosenblatt method works well to calibrate a mapping function, which we tuned on validation data, to improve expected calibration error (ECE) of a confidence score [20].

Others are wary of relying on confidence predictions, especially if it is shown that expected calibration error is difficult to improve, or if future datasets may have a distribution shift [21]. Another study showed very modest gains after calibrating for confidence with little significance on classification performance [22]. In our study, ECE was improved reliably for each model, and classification performance shows very significant performance when comparing the most confident samples versus the least confident samples.

In this study, we evaluate two significant procedures to improve overall performance of clinical evaluation of prostate pathology. In the first procedure, we improve labels on training data for improved classification of prostate primary Gleason grading. We applied the RANSAC technique to both prune or relabel a proportion of the training samples, demonstrating the effectiveness and importance of training data refinement. As a second significant procedure, we calibrated our output classification probabilities (or confidence) to determine thresholds to filter ambiguous samples with weak predictions. Together these two procedures improve performance over 10% across all metrics. Our results are consistent with other recent reports and ablation studies that have boosted performance through confidence filtering either on the training data [23–25] or after prediction and before final classification [26, 27].

Our experiments included several well-studied optimization techniques to improve performance as a series of ablation tests; the results are critically analyzed by repeating the tests multiple times and computing statistical significance [28]. The most significant improvements were gleaned from data sanitizing on both the training side and inference (or prediction) side; the former by label correction and sample rejection and the latter by using calibrated confidence on inferred label samples to ignore ambiguous examples. Fig 1 shows a rough model of the processes involved in both training and inference and will be discussed in detail in the next section.

**Fig 1.**
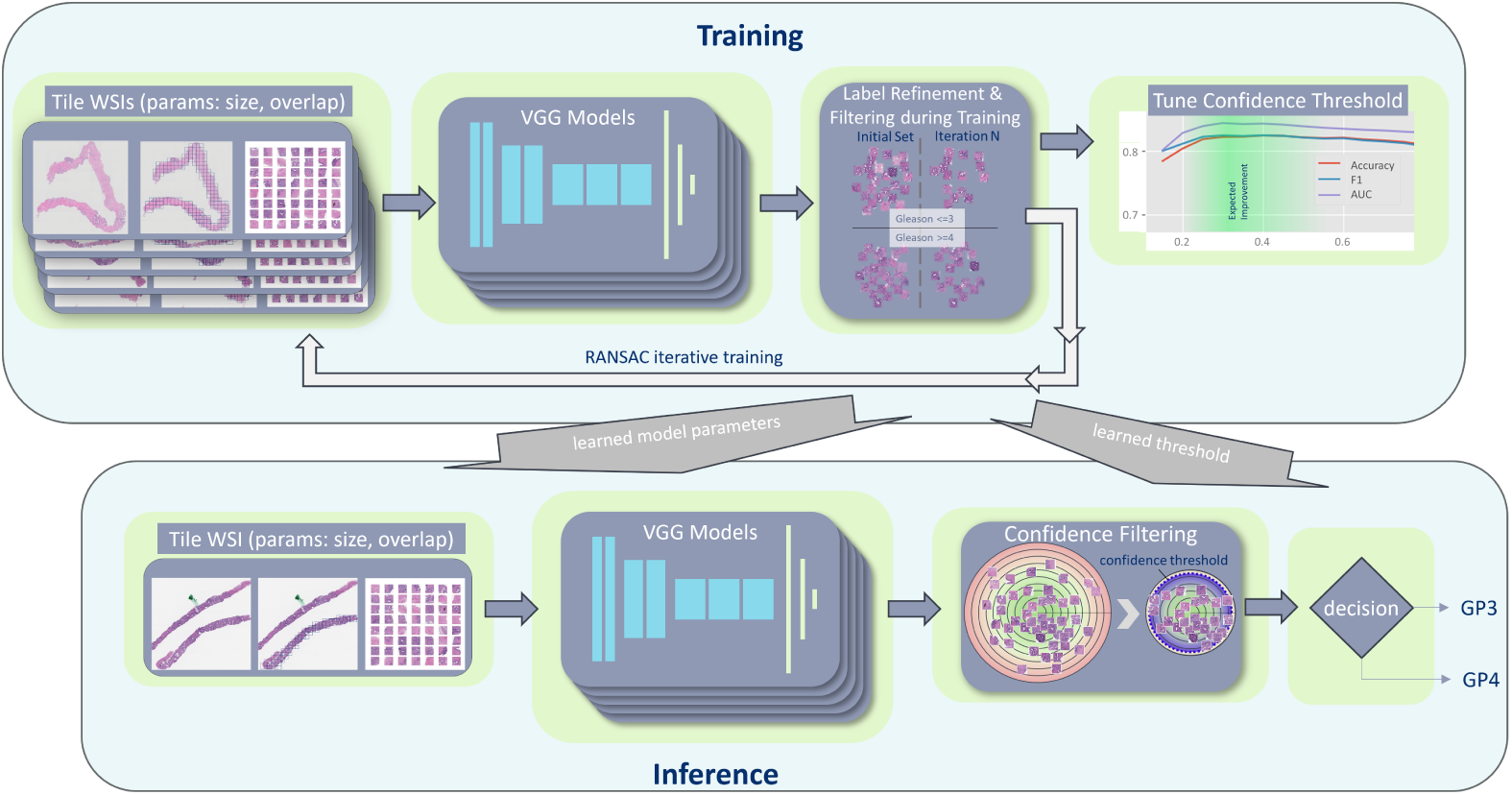
Training and Inference performance improves with data sanitization.

## Materials and methods

### Data Cohort

We retrospectively collected patient data (P=58) who underwent targeted biopsies (N=110) to evaluate prostate disease. Pathology slides were obtained for a cohort that had diagnosis and treatment at Moffitt in the years 2018-2019. The retrospective study was approved by the institutional review board (IRB) at Moffitt/University of South Florida. Informed patient consent was waived for retrospective research efforts for samples collected under the institutional protocol (Total Cancer Care^®^). The clinical records of the patients were obtained and pathological whole slides were digitized at high resolution (20x, 0.504*µm*/pixel) using an Aperio^®^ slide scanner. The research pathologist assessed the WSI and marked contiguous regions to provide a score following Gleason criteria, limited to the prominent pattern (primary Gleason score) at the region level. The digitized slides and derivative tiles were used in this study from 01/03/2025 to 31/08/2025 (March-August) to compile these research results.

We systematically divided the marked regions into smaller sections or tiles that are representative of the whole slide images and assigned the labels provided at the regions to the smaller tiles. The primary scores for these tiles/regions follow the ranges of patterns (benign, Gleason score 3, 4, or 5). Gleason 4 labels were not discriminated between cribriform and non-cribriform variants of malignancy, despite its role in aggressive disease prognosis [3, 29]. Despite collecting benign and Gleason 5 samples, their under representation excluded them from the tests in our study. The converged data cohort consisted of 3311 Gleason-pattern 3 (GP3) and 2909 Gleason-pattern 4 (GP4) labeled glandular patches. Fig 2 shows a random selection of primary GP3 (top panel) and primary GP4 at the bottom.

**Fig 2.**
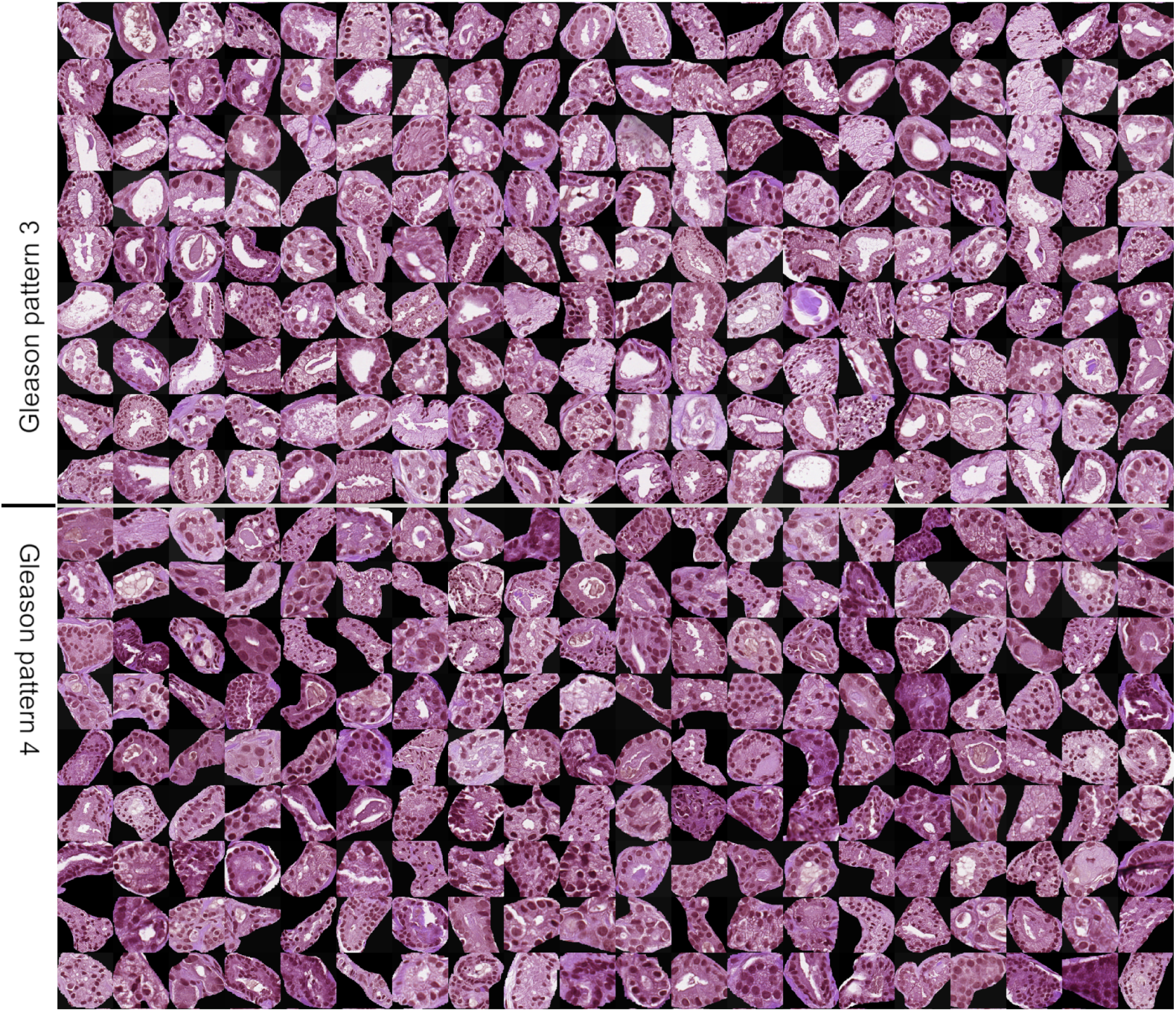
Random selection of Moffitt GP3 and GP4 indicated glands.

### Data split for DL modeling

We organized the data into seven randomized experiments (80/20 training/test ratio), with a total of 110 patient biopsies; each of the shuffled experiments had 88 training and 22 test WSIs. The seven training sets were further split into a five-fold cross-validation with each fold comprising 80/20% training/validation and evenly stratified. The CV folds were used to compute validation metrics for tuning ML models and later for creating ensembles of models. DL training data was shuffled between iterative runs and truncated (as necessary) to have a balanced representation of the labeled classes. The final metrics were computed by bootstrapping the data with both groups (GP3/GP4) in perfect proportion, and reported results are averaged over bootstrapped samples (100 times). Depending on imbalance, bootstrapped metrics were sampled from up to 75% of the holdout data. Cross-validated data splits are discussed in more detail in Supplemental **??**.

### Deep Models

We used tile-level data with glandular structures derived from whole slide images, and these patch images were preprocessed to standardize the data before sending them to a deep learner (DL). Our DL was a modified version of a VGG-16 architecture, limited to 3 groups of CNN filtering (or 7 total CNN stages) and two fully-connected layers replacing the 3-layer top of the standard VGG-16 [30, 31]; early experiments showed a full VGG-16 to easily over train and for generalization support to suffer. Thus, our optimized architecture was effectively a VGG-9 CNN (or a VGG-15 if non-optimized, using all CNN stages but a simplified top layer).

Stochastic gradient descent was used as the optimizer with learning rate scheduling following an exponentially decreasing cyclic cosine schedule. Training data patches whose predicted label after training disagreed with GT more than 80% of the time were relabeled, and the next 20% of patches that disagreed more than 60% of the time were removed from the training set. Results were improved by combining classifiers across 5 CV folds.

### Ablation Studies

DL ablation studies were nominally performed with the TensorFlow/Keras framework on a VGG-9 or VGG-15 CNN. The network’s initial weights (CNN layers) were transfer-learned from model trained on ImageNet data. All studies were performed on an NVIDIA^®^ DGX™platform with up to eight A100 GPUs.

The image patches were preprocessed to standardize the glandular data, which involved: resizing (to 300x300 pixels), converting to gray level (0-255), and masking of epithelial glands. The preprocessed data was also augmented via flips and rotations while training the deep learning network to improve generalization. Other hyper parameters (learning rate, momentum, batch size, dropout, etc.) were tuned to support validation data performance and following modern convention. Relevant source code and scripts may be referenced for additional detail at https://github.com/rfogarty/ConfidenceFiltering.

### Classifier Optimizations

Two DL model optimizations are important enough to warrant discussion: data sanitization and an ensemble of models. Both techniques provided significant gains in performance and will be briefly discussed in our results. We used the random sample and consensus (RANSAC) approach, previously studied and reported in [13], to prune difficult samples from the training set [14]. We also applied label-flipping when samples rarely compared correctly to the ground-truth label. Further details of the algorithm and thresholds used to label-flip or prune samples are discussed in Supplemental **??**. Ensembles were used by voting on inference decisions across 5-fold cross-validated models. Similar gains were made using ensembles from the best 5 models using snapshot ensemble approach.

We performed many other ablation studies, which include preprocessing of image patches (graying, cropping, masking) and deep network tuning (model pruning, exponentially-decreasing learning rate scheduling) - each only provided small improvements and are not discussed in detail in this paper. Several of these hyperparameter, neural network architecture, and data preprocessing steps were ablated to study their effect. Because their effect was small, the results of those tests are not included here for brevity and are reserved for publication at a later time.

### Confidence Filtering

In a sample discrimination context with binary classes, we can order samples based on confidence and ignore predictions if alternate observations of a sample provide stronger evidence of a state or classification. We propose to remove those predictions with low confidence to improve decisions. The pseudo-confidence produced by a discriminate function (such as the output of the sigmoid of a binary DL classifier) can be used to derive a confidence on each sample’s decision. Whereby, the least confident samples are discarded when considering the larger picture or an overall classification decision.

To measure confidence, the output of a DL classifier can be mapped to a pseudo-confidence (*p̂_i_*) and then calibrated towards a true confidence (*q̂_i_*). In an ideally calibrated system, a group of samples (say *n* samples) between two thresholds should each produce a pseudo-confidence that closely ties to the system’s measured accuracy. E.g., if we were to take *n* samples produced with a pseudo-confidence of between 0.9 and 1.0, then in the average we expect *>* 90% of these *n* inferences to classify correctly; we presume a frequentist interpretation of model behavior; we can both measure calibration error and correct for it by remapping to measured performance [18, 32, 33].

A mapping function for calibrated confidence is estimated using cross-validation data from five-fold results (see below). Prior works have produced various methods for estimating this calibration function, such as histogram binning, isotonic regression, and Platt scaling [33]. Each method estimates a mapping function between the true confidence and the pseudo-confidence derived from a model’s probabilistic output. We first define a pseudo-confidence, *P̂*, relative to the DL model’s (sigmoidal) binary output *Ŷ*. The *i^th^* pseudo-confidence is:

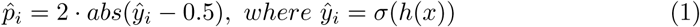

The goal is to map *P̂* → *Q̂* = *p*, or a calibrated confidence, so that

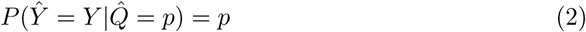

or near perfect calibration. A generalization to histogram binning is kernel density estimation also termed the Parzen-Rozenblatt window method [20]. A closer approximation to the true underlying confidence distribution is achieved by convolving a correctness array *C* (boolean) with a kernel function. Before convolving the array *C*, it is first sorted in order of pseudo-confidence *P̂*. The result of the kernel convolution estimates the cumulative density function. The kernel chosen was a one-sided falling exponential function of the form:

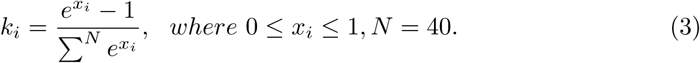

The kernel window was empirically adjusted to perform well with a length of *N* = 40. The KDE algorithm computed for each model independently is as follows:

Step 1. Compute inference results on validation samples,

Step 2. Create boolean array *C* (correct classification for each inference),

Step 3. Convert each sample’s DL test statistic output (sigmoid in binary case) to *p̂_i_*,

Step 4. Sort the boolean array *C* by lowest to highest *p̂_i_*, Step 5. Convolve sorted *C* with kernel function to produce *Q̂*,

Step 6. Sort array *Q̂* to ensure it is monotonically increasing *Q̂_corrected_*, Step 7. Ensure remap array *Q̂_corrected_* range [0.0-1.0].

Note that Step 6 is a small accommodation to enforce monotonicity and allow for an unambiguous mapping function. The algorithm trained on validation data and tested on holdout data reduced expected calibration error (ECE) from 22.6% down to 10.2%, or from 20.6% down to 8.6% when ensemble scoring. ECE was computed using the following standard algorithm:

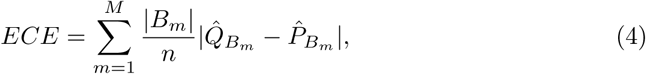

where *P̂_B_m__* is average pseudo-confidence in bin (*B_m_*), and *Q̂_Bm_* is measured accuracy within that same bin (*B_m_*), and M is a reasonable number of bins to test, which was set to 20 in our case.

Alternatives such as the histogram binning technique exhibited slightly lower calibration error than kernel density estimation (KDE), but bins were not as numerically stable and the resultant mapping function was not as parsimonious as the KDE approach. Results for histogram binning method are shown in more detail in the Supplemental **??**.

Applying our KDE algorithm independently for each trained model, Fig 3 shows the scatter plot for 35 calibrated models showing uncalibrated or pseudo-confidence *P̂* along the x-axis, and calibrated confidence *Q̂* along the y-axis. Each point in the plot is a single validation sample (from a training-validation set) plotted with coordinates *p̂_i_* and *q̂_i_*, where *q̂_i_* ≃ p is an estimate of a sample’s true confidence. Because we do not have infinite samples to use for calibration, the remapping function is a piecewise monotonically increasing mapping function, whose smoothness depends upon the number of validation samples.

**Fig 3.**
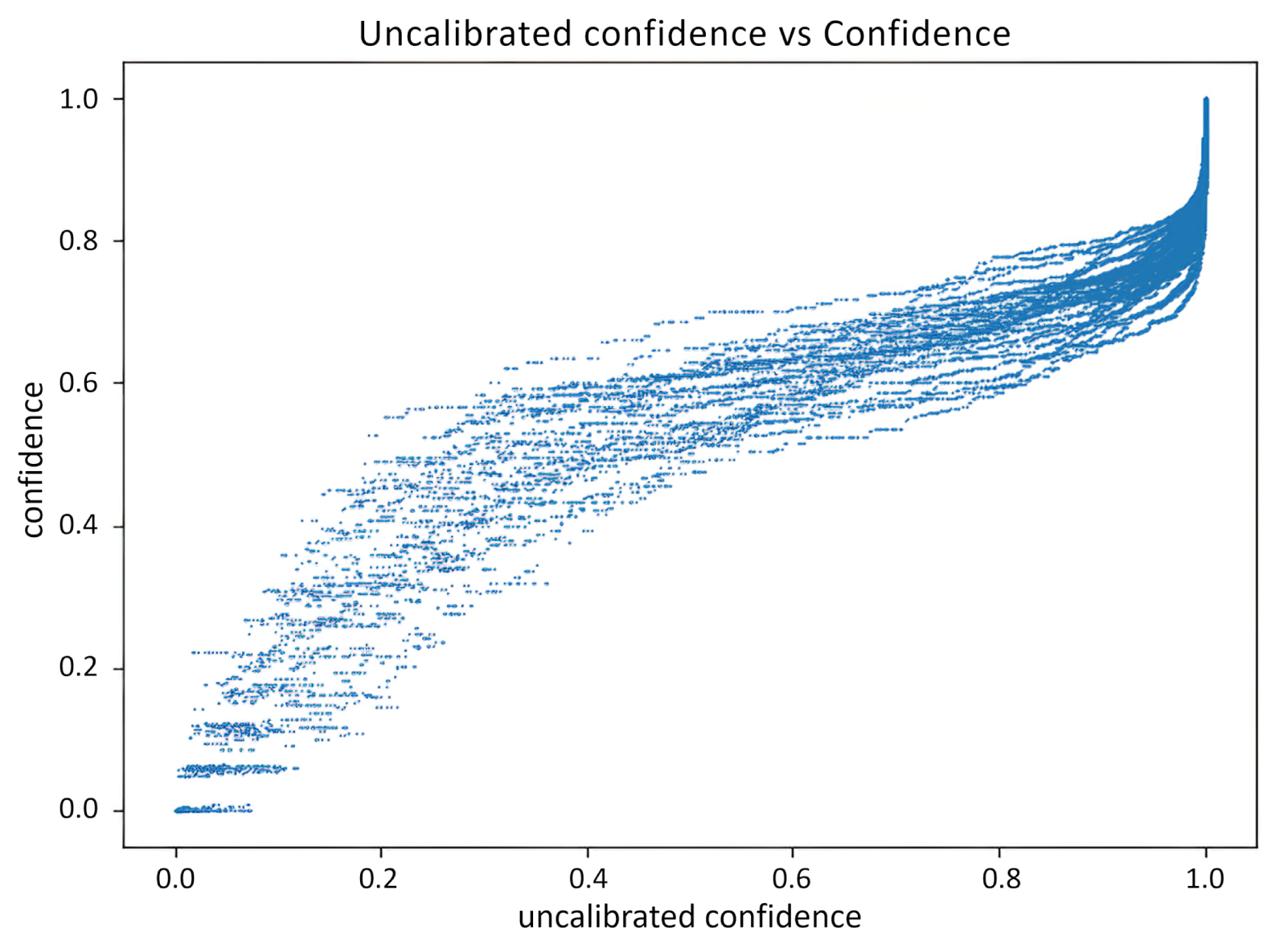
Calibrating confidence for 35 (7 experiments * 5 CV folds) VGG9 trained models.

After calibrating confidence, performance is improved by throwing away samples with low-confidence. Fig 4 shows an algorithm for estimating a good confidence threshold or a sampling fraction that produces improved results. Fig 5 shows the related algorithm for measuring performance on holdout data and resultant summary statistics, which ultimately were determined by evaluating the 35% highest confidence samples, discussed in further detail below.

**Fig 4.**
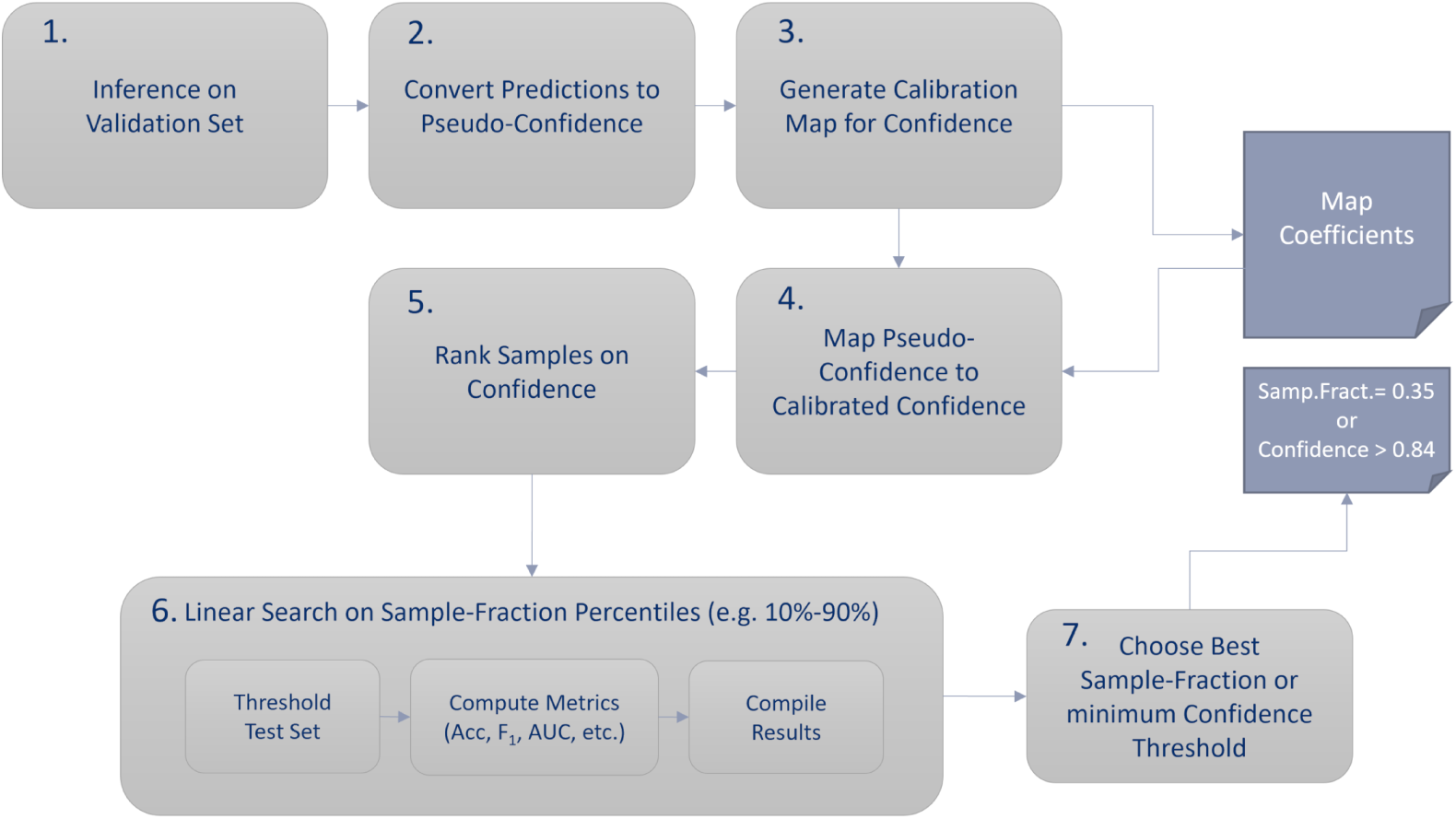
Confidence Threshold Search Algorithm.

**Fig 5.**
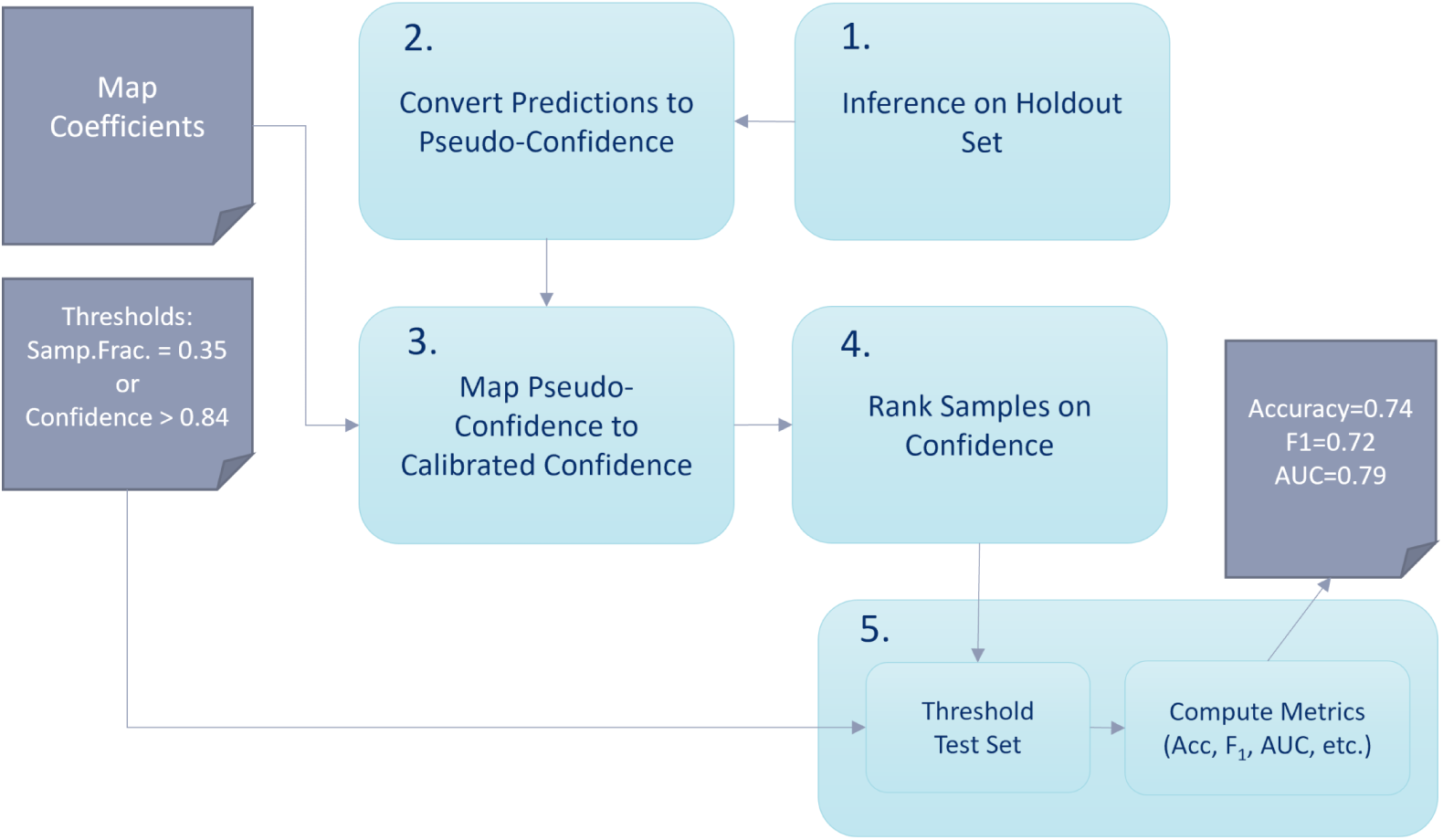
Testing Performance with Confidence Threshold.

## Results

We used deep learning architectures (VGG) initialized with ImageNet-trained weights and fine-tuned on our prostate pathology cohort. We report results averaged over multiple (reshuffled) experiments. Final metrics were derived from holdout sets unseen during training. The holdout sets were grouped such that patches from any one whole slide image were exclusive to the training/validation set or to the holdout set.

The study curated over 6,000 patches, with 3311 primary GP3 and 2909 primary GP4 glandular patches labeled by the research pathologist. In total, 35 DL models (7 reshuffled experiments with 5-CV folds each) were trained for each ablation test or study. The accuracy, F_1_ and AUC metrics reported are average and standard deviation over all experiments.

### Ensembles’ Significance

Ensemble results were computed on a VGG-9 architecture and over 16 separate experiments studying various micro-optimizations (grayscale versus RGB, 4 patch resizing strategies, and masking on/off), with each of the 16 experiments showing ensemble improvement (though not shown here). We found creating ensembles over the five-fold CV models shows a very strong effect in increasing generalization performance (see Fig 6). The boxplots qualitatively show a significant gain in accuracy, F_1_ and AUC scores, and tighter variances. The Cohen’s d effect size along with the Wilcoxon ranksum p-values shown in the x-axis labels provide a quantitatively strong significance across all metrics when comparing non-ensemble results on the left and ensemble models on the right. In total, metrics were computed for 35 single models and 7 ensembled models and repeated for 16 ablation experiments, which led to 560 simple models and 112 ensemble models; from these results we drew 50 samples in each category (without replacement) and computed significance over 1000 times. Due to the strong positive effect of ensembles and a 3-4% increase in all performance metrics, all additional experiments assume the ensemble optimization unless otherwise stated.

**Fig 6.**
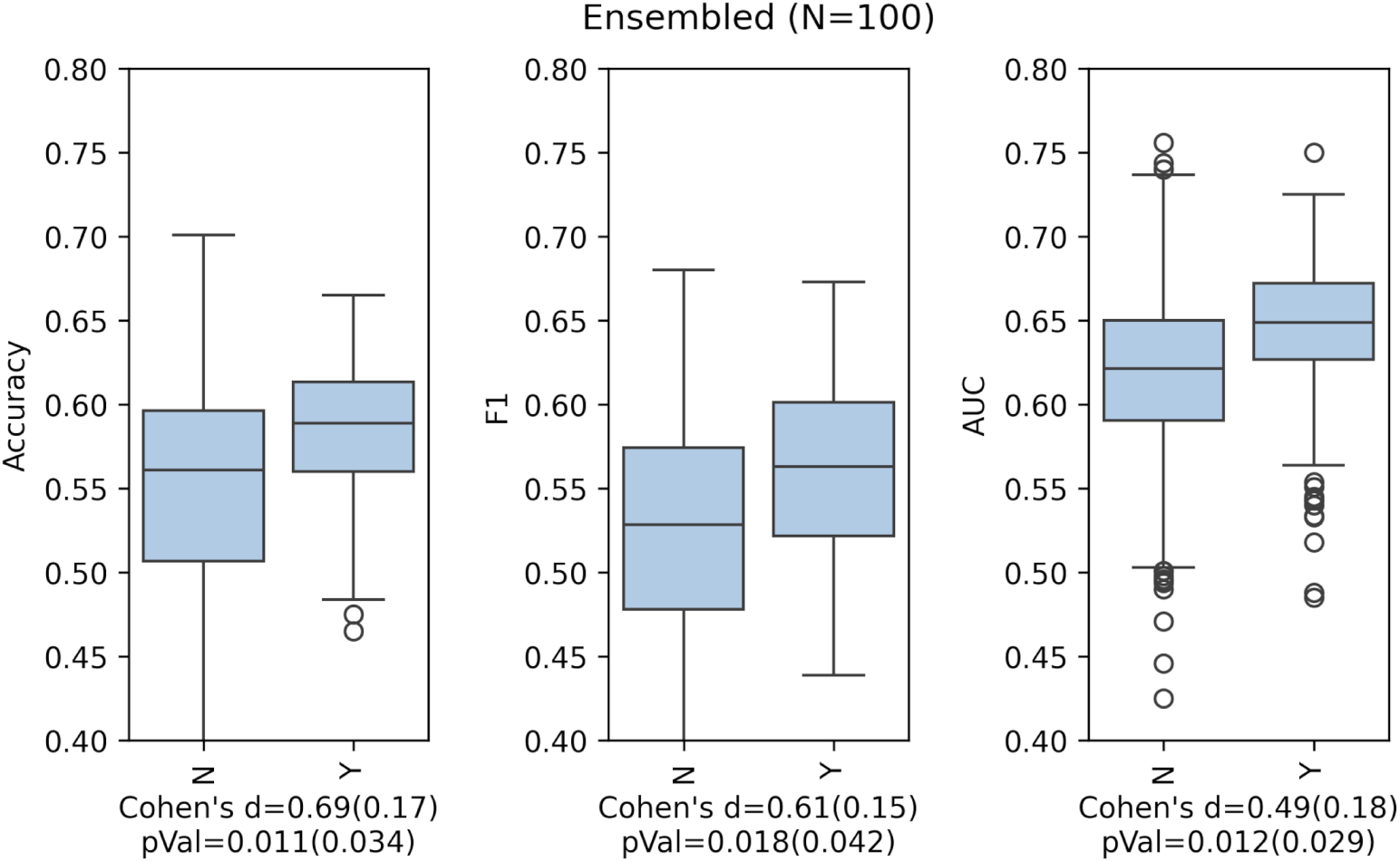
Ensembling across CV folds improved performance significantly.

### Training Data Refinement

We iteratively improved our training data by discriminating the samples into Gleason pattern groups and verifying improvement in classification performance. Data sanitization included pruning ambiguous samples or flipping labels when patches very rarely matched ground truth.

Applying data sanitization and ensemble optimizations together achieved a 0.68 accuracy, 0.66 F_1_ and 0.74 AUC for a DL when discriminating glandular labels between GP3 and GP4 samples. The results very closely matched our previously published performance of similarly-sized patches derived from a University of Miami data cohort as shown in Table 1 [13].

**Table 1.** Deep learning (CNN) performance on Moffitt and Miami.

| | Accuracy | $F_1$ | AUC |
| --- | --- | --- | --- |
| Moffitt mean (stddev) | 0.68 (0.04) | 0.66 (0.06) | 0.74 (0.04) |
| Miami mean | 0.67 | 0.66 | 0.71 |

In this ablation test, a random sampling and consensus (RANSAC) approach was used to identify the lowest performing training/validation samples. Incorrectly inferenced training/validation samples with more than 80% error were flipped in label (between GP3 and GP4) and corrected in the training set for a subsequent fine-tuning period. The next least performing samples (incorrectly predicted more than 60% but less than 80%) were pruned from the training and validation sets. The results of label refinement were computed on the complete and *unmodified* holdout sets and based strictly on GT labels. The metrics in the boxplots of Fig 7 include 2 columns *Original* and *Updated*. The metrics in *Original* columns are derived from 35 models trained on the complete training with GT labels, while the metrics in *Updated* columns were results from 35 models trained on a pruned set and refined labels. Significance tests are computed over the N=70 total models from each group. As will be shown, these results are improved further through confidence filtering.

**Fig 7.**
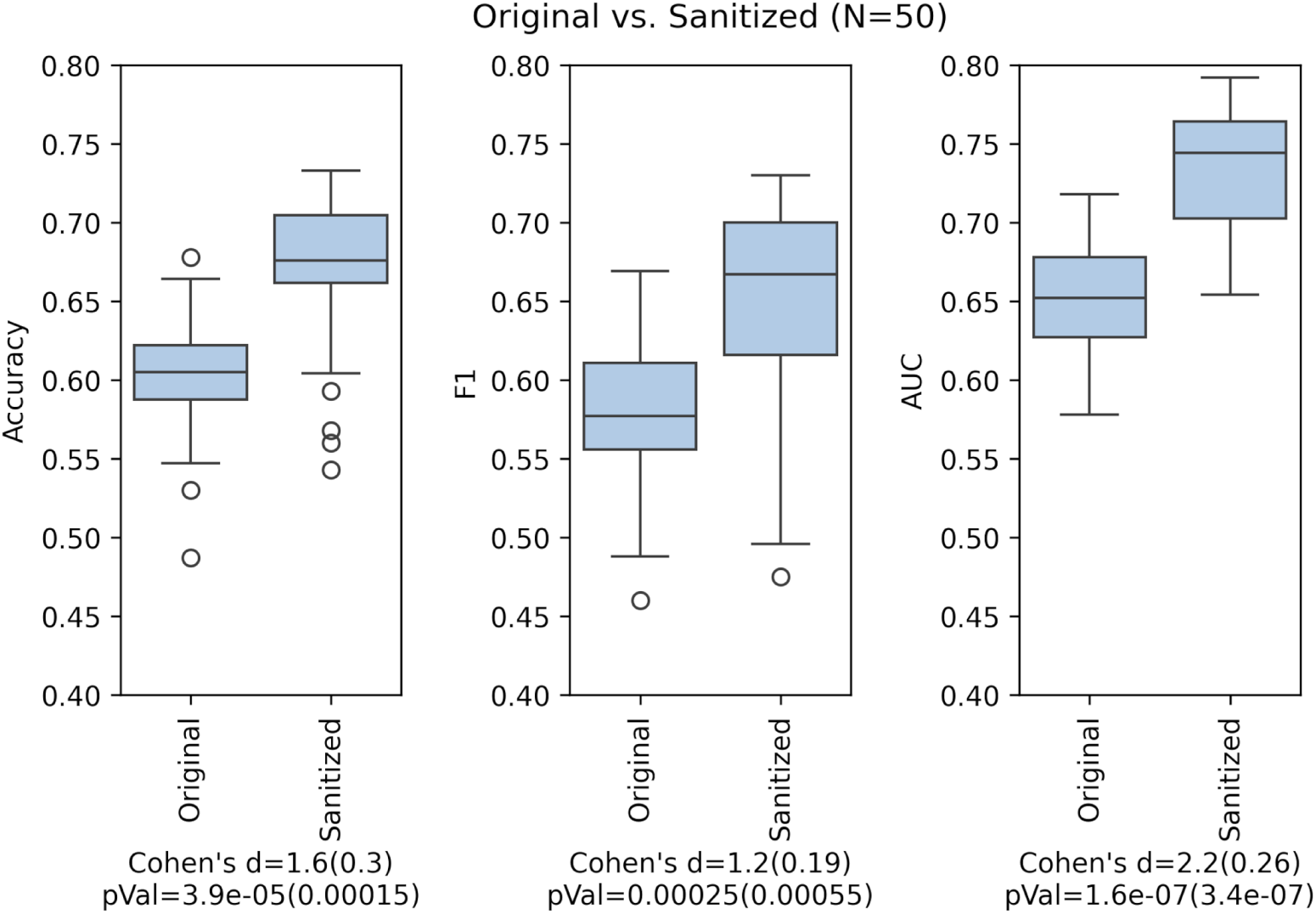
Performance improves significantly with training data sanitizing.

### Confidence Filtering

The results of confidence filtering on our holdout sets (filtering sample inferences with lowest confidence) improved results more than 5% demonstrating an accuracy of 0.74, F_1_ of 0.72 and AUC of 0.79.

To determine if a filter threshold or sample fraction expects improved performance, we first ranked validation samples according to pseudo-confidence *P̂*. We ranked all inferences according to their *p̂_i_*, and measured performance at filtering thresholds incremented by 5% starting at 0% and ending at 85%. Fig 8 shows that validation performance slowly improved in all metrics of accuracy, F_1_ score and AUC, with an eventual peak 5-10% improvement over the baseline, which is nominally the result at 100% *sample-fraction* or no filtering. We will show that peak-performance on the validation set was at a filter threshold of ∼ 65% of the samples (or maintaining 35% *sample-fraction*). Beyond 75% filtering, performance began to taper off rapidly.

**Fig 8.**
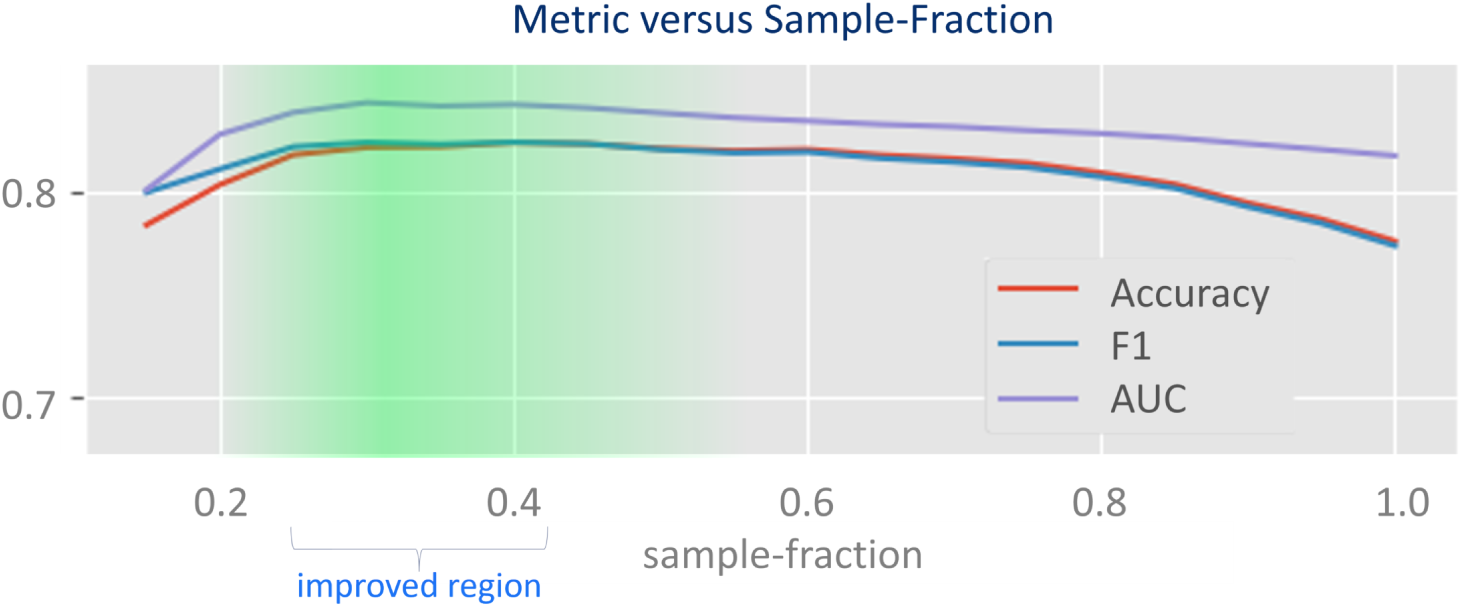
Accuracy, F_1_, and AUC versus *sample-fraction* observed on validation data.

We observe the validation data in detail to show the effect of filtering on confidence for our discriminant classifier (the VGG-9 CNN) to forecast utility in the general case. Looking at the numbers in Table 2 shows confidence-filtered accuracy, F_1_ and AUC metrics improving at each 5% threshold tested until about the 0.70 threshold. The improvement in metrics demonstrates a clear opportunity to discard or ignore low-confidence samples to improve our overall performance.

**Table 2.**
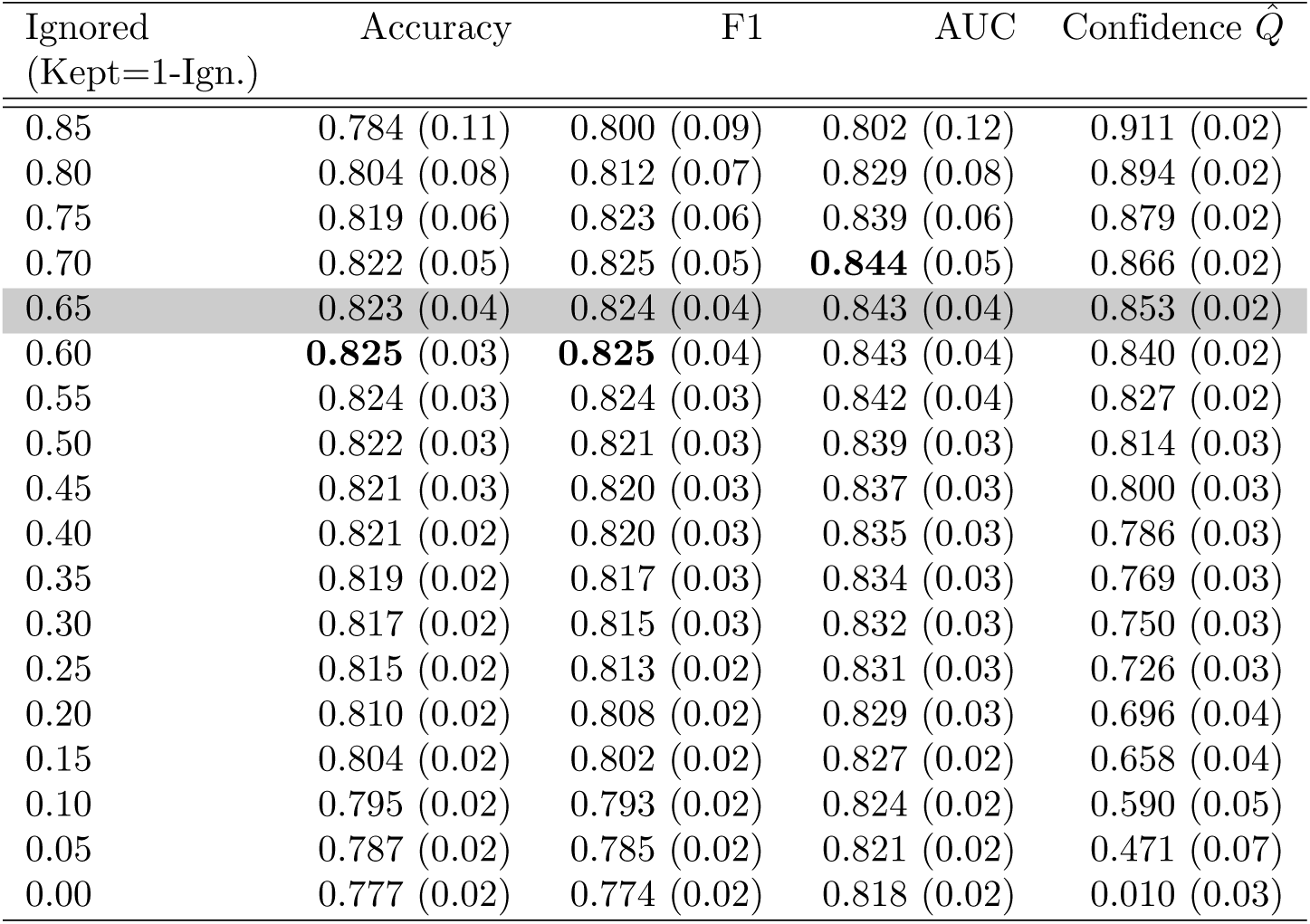
VGG-9 CNN validation performance versus *Sample-fraction*.

| Ignored<br>(Kept=1-Ign.) | Accuracy | F1 | AUC | Confidence $\hat{Q}$ |
| --- | --- | --- | --- | --- |
| 0.85 | 0.784 (0.11) | 0.800 (0.09) | 0.802 (0.12) | 0.911 (0.02) |
| 0.80 | 0.804 (0.08) | 0.812 (0.07) | 0.829 (0.08) | 0.894 (0.02) |
| 0.75 | 0.819 (0.06) | 0.823 (0.06) | 0.839 (0.06) | 0.879 (0.02) |
| 0.70 | 0.822 (0.05) | 0.825 (0.05) | **0.844** (0.05) | 0.866 (0.02) |
| 0.65 | 0.823 (0.04) | 0.824 (0.04) | 0.843 (0.04) | 0.853 (0.02) |
| 0.60 | **0.825** (0.03) | **0.825** (0.04) | 0.843 (0.04) | 0.840 (0.02) |
| 0.55 | 0.824 (0.03) | 0.824 (0.03) | 0.842 (0.04) | 0.827 (0.02) |
| 0.50 | 0.822 (0.03) | 0.821 (0.03) | 0.839 (0.03) | 0.814 (0.03) |
| 0.45 | 0.821 (0.03) | 0.820 (0.03) | 0.837 (0.03) | 0.800 (0.03) |
| 0.40 | 0.821 (0.02) | 0.820 (0.03) | 0.835 (0.03) | 0.786 (0.03) |
| 0.35 | 0.819 (0.02) | 0.817 (0.03) | 0.834 (0.03) | 0.769 (0.03) |
| 0.30 | 0.817 (0.02) | 0.815 (0.03) | 0.832 (0.03) | 0.750 (0.03) |
| 0.25 | 0.815 (0.02) | 0.813 (0.02) | 0.831 (0.03) | 0.726 (0.03) |
| 0.20 | 0.810 (0.02) | 0.808 (0.02) | 0.829 (0.03) | 0.696 (0.04) |
| 0.15 | 0.804 (0.02) | 0.802 (0.02) | 0.827 (0.02) | 0.658 (0.04) |
| 0.10 | 0.795 (0.02) | 0.793 (0.02) | 0.824 (0.02) | 0.590 (0.05) |
| 0.05 | 0.787 (0.02) | 0.785 (0.02) | 0.821 (0.02) | 0.471 (0.07) |
| 0.00 | 0.777 (0.02) | 0.774 (0.02) | 0.818 (0.02) | 0.010 (0.03) |

Improved performance is achieved when *sample-fraction* is in the range of 30-40%, which corresponds to a minimum calibrated confidence of ∼ 0.85 at around the 0.35 *sample-fraction* threshold. So, one approach is to use calibrated confidence levels *q̂_i_* ≥ 0.85 to discard low-performing samples on future unseen data. To use percentile *sample-fraction*, we only need to rank and sort samples of an unseen subject and prune the 65% lowest performing samples as a reasonable threshold as shown in the VGG-9 table line highlighted in gray; this latter approach was decided to be the best method before testing on holdout data to guarantee a reasonable sample size for any subject. Either approach can be used as a litmus test for the quality of inferenced samples or as a secondary approach to filter low-confidence samples.

After making the determination to use the 35th percentile (ignoring lowest ∼ 2*/*3 of inferences ranked by confidence) as a reasonable threshold on unseen data, we checked how we performed in actual holdout test performance. Results for all sample-fraction thresholds are shown in detail in the Supplemental **??**.

Summarizing the results of holdout testing with our chosen 65% threshold (i.e. keeping highest 35%) is shown in Table 3. Performance is improved over 5% for the DL demonstrating an accuracy of 0.74, F_1_ of 0.72 and AUC of 0.79, and improving prior work classifying prostate pathology glandular patches in the 40-100 micron range. The tradeoff for ignoring low-confidence samples is higher inference variance; the standard deviation is doubled across all performance metrics.

**Table 3.** Refined VGG9 performance ignoring least (65%) confident samples.

| Filter | Accuracy | F1 | AUC |
| --- | --- | --- | --- |
| confidence-filtered | **0.74**(0.08) | **0.72**(0.12) | **0.79**(0.08) |
| without filtering | 0.68 (0.04) | 0.66 (0.06) | 0.74 (0.04) |

### Total Optimization Effect

Finally, in this study, we compared our optimized network with confidence filtering to a network with the full VGG-16 CNN layers (5 stage) to serve as a näıve baseline. The baseline network utilized the AdamW optimizer (default settings), performed basic image preprocessing (simple rescale, full-color patches, no gland masking), and no data sanitization (label flipping or pruning of training data). In both this baseline case and our optimized architecture, the models were trained for over 500 epochs or generally until we observed validation loss convergence.

Here we present the optimal case with ensembles versus the baseline unoptimized case without ensembles. As shown in Fig 9, boxplots are drawn for accuracy, F_1_ and AUC for 7 ensembled models in the optimized case and 35 un-ensembled models in the baseline unoptimized case (5-folds x 7 repeated tests), for a total sample size of *N* = 42. The Supplemental **??** presents the significance comparisons when the optimized models and the näıve baseline are both without ensembles and both are with ensembles; regardless of the approach, the optimized models show much higher performance. Taken in concert, the optimizations used in this study account for a much improved result over the näıve baseline and the p-values computed with the Wilcoxon rank-sum test shown below each metric pair report high significance.

**Fig 9.**
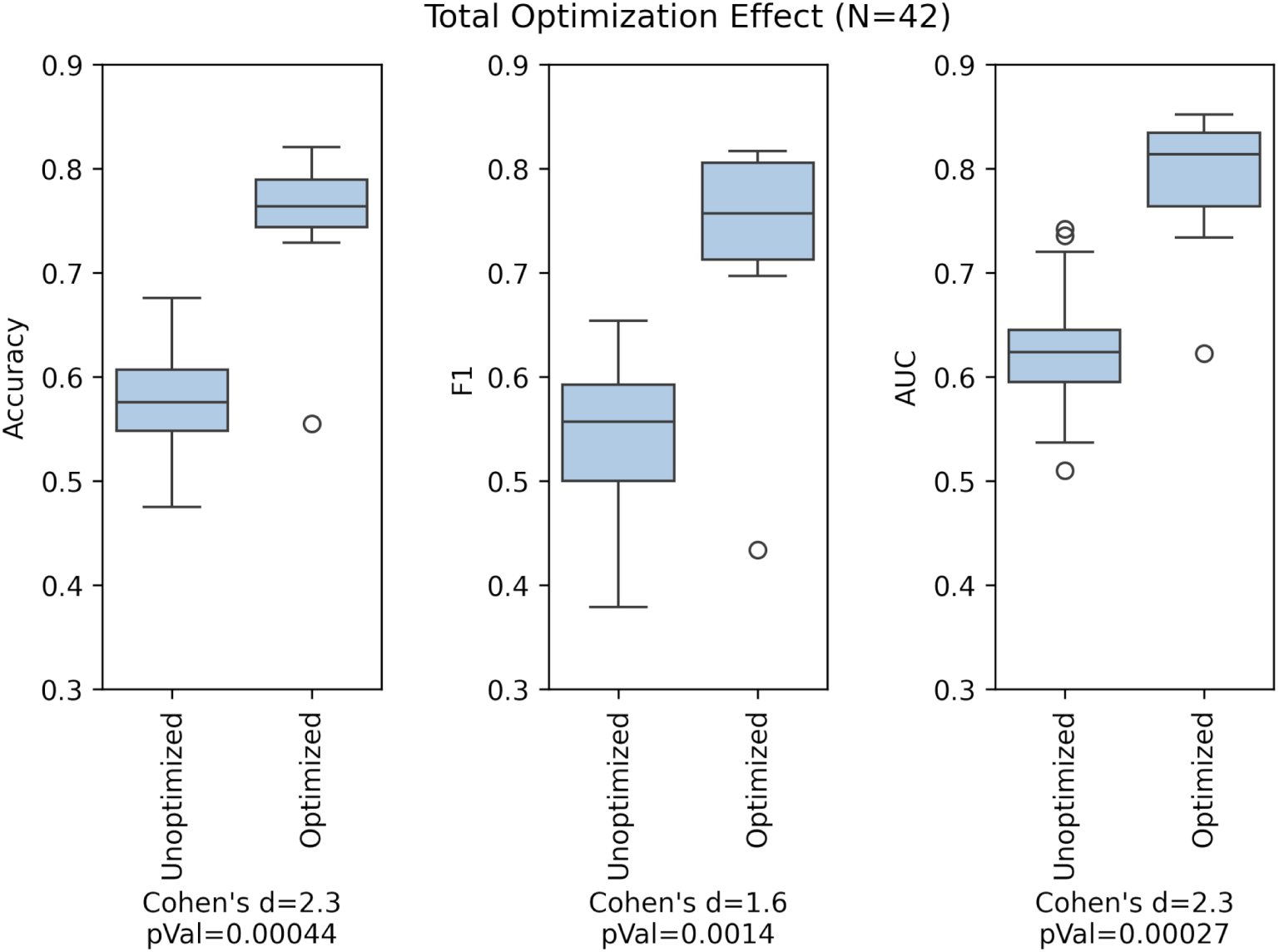
Total performance improvement versus näıve baseline.

## Discussion

Histopathological grading of prostate samples is challenging, and expert opinions still differ with a reported low concordance [2]. In this study, we focus on assembling a cohort of exemplars of whole slide images with smaller patches of uniform glandular regions with labels derived at the tile level from a clinical expert. We built a statistical procedure, called ‘confidence filtering’, adapted for deep models, to grade the samples, which allows us to focus on high-confidence samples to improve deep models’ discrimination ability. The hypothesis assumes inference error is a function of pseudo-confidence, therefore, if we eliminate lower-confidence samples, measured classification performance will improve.

Training data is improved by removing samples that classify incorrectly across a diverse set of models. We find further improvements in trained models by flipping labels on samples that appear to be egregiously incorrect (or incorrect with high-significance).

Gleason patterns on small glandular features are a challenging problem for expert histopathologists and AI algorithms alike. At such a fine-level, we require careful tuning and exploration of model selection and parameters to achieve practical performance. Proper tile preparation (grayscale, resize by cropping, and masking), model simplification, and robust learning rate scheduling provides measurable but minor improvement so was largely left out of the discussion. More significant improvements are found by ensembling, label refinement, and ignoring low-confidence inferences.

Disregarding all of these techniques, it is difficult to achieve discrimination of Gleason pattern 3 and 4 on 40-100 micron patches much above chance as was shown on a näıve baseline, which had mean accuracy and F_1_ around 57%. The major optimizations improved this performance by almost 20%. As others have reported sample labeling biases influence the model performance [17]. There are several prior works that proposed selective classification in the context of better uncertainty quantification [19, 26, 27, 32, 34, 35] and data sanitizing in the context of classifier training [15, 23–25, 36].

DL and ML has shown to be effective at determining prostate cancer (PCa) from non-cancer with very high accuracy but is more challenging in determining degree of malignancy between low-grade and high-grade prostate cancer or neighboring Gleason patterns. Butt et al reported high performance on the SICAPv2 dataset with 512x512 images at 10x resolution by applying multi-labels to individual patches; though results cannot be compared directly since individual patch-level performance is not reported [6]. Duenweg et al classified patches with a bagged ensemble ML and ResNet-101 DL. Similar to our study, the authors classified individually-labeled patches of 1024x1024 (∼ 1020*µm*^2^) in size. Their results of comparing high-grade (HG, Gleason 4 or 5) and low-grade (LG, Gleason 3) cancer is comparable to our GP3 versus GP4 study. In their best case with a ResNet-101, they achieved an overall accuracy of 0.72 [3], while our result achieves 0.74 accuracy after confidence filtering. Our result does not include Gleason pattern 5 samples, which in theory would improve our result further since GP3 and GP5 Gleason patterns have more differentiation than GP3 to GP4 samples.

Our results improved on a previous study on multi-institutional data [13] but the results in this study are computed using the stronger holdout test methodology (versus Monte-Carlo CV used in prior studies). In Fig 7, we showed boxplots to demonstrate qualitatively that refining the training/validation sets through pruning and relabeling significantly improves holdout performance; holdout data is strictly based on original GT. The strong performance improvement in label flipping supports the need for clean data prior to clinical application and has practical applications across areas that depend on expert opinions. Aleatoric noise is evident in most clinical studies that are often dependent on expert opinion and often influenced by their training [17]. It is evident that the Gleason pattern in prostate pathology is known to have high variability due to the complexity in the interpretation of glandular structures, coupled with limitations of specimen sampling [6].

If DL classification is performed over an entire WSI, the output decision is often too abstract to generate an evidence-based result and is vulnerable to confounding factors. Explainable AI (XAI) methods may someday shed light and provide further evidence on DL behavior, but a feasible approach is to train DLs to look and classify smaller regions. The deep-classifiers presented in this study perform at a much finer-level than those published in competing works, which makes classification more challenging. These classifiers can be used in a suite of decision support system tools to augment drill down interrogation of WSI classification, or to build support for an overall Gleason grade decision.

## Conclusion

The study results demonstrate the efficacy of using deep models to classify glandular patches at a fine scale. Confidence filtering on properly calibrated DL models demonstrated a performance increase and is appropriate in this modality with noisy labels, which is a common problem in most human expert-driven studies. Our study finds that glandular patches of prostate histopathology improve the discrimination of pathological grades using confidence filtering. We also find that ensembles of models and data sanitation (label flipping and pruning) along with confidence filtering have the largest impacts in designing a system to classify primary Gleason pattern at the gland level.

Evidence at this low level can operate in a larger suite of decision support system tools to augment and reify expert pathology decisions and improve concordance among clinicians. We envision a decision support system for pathologists that improves performance by confidence filtering either by rank sorting, calibrated confidence thresholding, or both. When using an ensemble of models, calibrated confidence from each prediction can be used to compute a discriminate average versus simple voting schemes. Additionally, clinicians can observe a subject’s array of calibrated confidences *Q̂* to make insights on the quality of the overall classification decision.

## Limitations

Current work is at the glandular level; we expect to leverage techniques developed in this research to improve classifiers that observe larger patches of the prostate specimen.

We did not attempt to optimize a clinical score over entire WSIs or patients as the data cohort was not evenly labeled across subjects - many patients had very few epithelial glands or regions labeled, with limited expert-level review.

## Data Availability

All data produced in the present study are available upon reasonable request to the authors.

## Acknowledgments

We would like to acknowledge our patients, care providers, and the voluntary contribution of biological samples to our institutional repository (Total Cancer Care), which has enabled us to conduct this study. We are grateful to Dr. Alexis Lopez and Joseph Johnson, along with their core team members, for providing research annotations and data preparation.

