## Supplemental Document for "Improving automated prostate pathological grading via confidence filtering"

### Confident Filtering: Supporting Information

#### S1 - Data Splits

Supplement Table 1 shows the number of validation samples in each CV fold, along with the number of holdout samples for each experiment (0-6). Note, the amount of training data for each CV fold isn't stated directly, but may be easily derived by summing the number of GP3/GP4 samples in CV folds outside the fold in question (roughly about 3800-4200 training patches depending on experiment).

**Supplement Table 1.** Samples per validation fold and holdout test sets (GP3/GP4)

| | Fold 0 | Fold 1 | Fold 2 | Fold 3 | Fold 4 | Holdout Test |
|---|---|---|---|---|---|---|
| Set 0 | 525/526 | 526/527 | 526/527 | 526/527 | 528/528 | 680/274 |
| Set 1 | 473/470 | 474/471 | 474/471 | 474/471 | 479/473 | 937/553 |
| Set 2 | 490/433 | 491/434 | 491/434 | 491/434 | 493/439 | 855/735 |
| Set 3 | 535/440 | 536/441 | 536/441 | 536/441 | 538/443 | 630/703 |
| Set 4 | 550/478 | 551/479 | 551/479 | 551/479 | 556/482 | 552/512 |
| Set 5 | 551/413 | 552/414 | 552/414 | 552/414 | 553/417 | 551/837 |
| Set 6 | 487/469 | 488/470 | 488/470 | 488/470 | 490/471 | 870/559 |

#### S2 - Data Sanitation

##### Training Label Refinement and Sample Pruning

Since studies have shown that Gleason scoring at the biopsy or specimen level has historically seen a low concordance for Gleason scoring of just 0.68 among expert urinary histopathologists [1], it is not unreasonable to assume that many small regions and glands are also incorrectly labeled when judgement is by a single pathologist. Furthermore, due to the difficulty in discriminating regions (much less glands) to a particular Gleason pattern (or degree of malignancy), and given the odds of occurrence for mislabeling being very high, models are improved by removing such samples, or flipping their labels in the training (and validation) sets for samples with such extraordinary chance of occurrence. Thus, sanitizing our data cohort was determined to be instrumental in improving Gleason pattern classification. To clean the Moffitt data set we used the random sampling and consensus (RANSAC) approach, whose technique is well-known to identify outliers in regression analysis. We applied the following steps of RANSAC to build a fliplist and blocklist:

1. Iterate RANSAC approach multiple times to refine sanitization rules:

   (a) Repeat for each of seven(7) randomized experiments:

   Step 1. Build multiple models derived from five-fold cross-validation 80/20 splits of training/validation data,

   Step 2. For each model training fold, by using cyclic-learning and snapshot ensemble, save the five(5) best performing models (by lowest validation loss) over all training epochs,

   Step 3. Inference on all validation and *training* samples for each of the 25 models; note models in most cases (20/25) have "seen" the data that is being tested,

   Step 4. For each sample count the number of correct classifications out of the 25 inferences,

   Step 5. Use reasonable heuristic to relabel samples that almost always classify incorrectly and add to "fliplist",

Step 6. Use additional heuristic to remove samples that may be ambiguous and add to "blocklist",

(b) Merge fliplists and blocklists from each of seven(7) experiments (eliminating duplicates),

2. Append to fliplists and blocklists for each reiteration (eliminating duplicates).

To determine a good heuristic for flipping labels or removing samples from the training/validation sets we make a couple of simplifying assumptions. First, we assume that our classifier models converge on a solution and operate with low epistemic error. Secondly, we assume that ground-truth data has more information than random noise and will eventually find a discriminating function that separates the 2 classes. Lastly, we note that we are not attempting to eliminate a constant bias that may be prevalent in ground truth labels. Rather, this bias will be preserved in the final discriminator. From the Binomial distribution equation in Eq 1, we may characterize the likelihood of mislabeling error of glandular structures. Supplement Table 2 presents the p-values or chance of occurrence for events with y equal to or lower successes in N=25 trials. Using the insight of these p-values, we can build reasonable heuristics for mislabeled samples and ambiguous samples. Note, we assume an equal chance probability for either event $p = 0.5$ (and its null hypothesis $q = p - 1$) in deriving this table; though it is difficult to determine this prior $p$ in this case, the assumption of $p = 0.5$ provides the upper limit on p-value estimates.

$$P(Y \leq y) = \sum_{i=0}^{y} \binom{N}{i} p^i (1-p)^{N-i}. \tag{1}$$

**Supplement Table 2.** Tail probabilities of Binomial Distribution, N=25, p=0.5

| Binomial $P(Y \leq y)$ | p-value |
|---|---|
| $y = 0$ | **2.98E-08** |
| $y = 1$ | **7.75E-07** |
| $y = 2$ | **9.72E-06** |
| $y = 3$ | **7.81E-05** |
| $y = 4$ | **4.55E-04** |
| $y = 5$ | 2.04E-03 |
| $y = 6$ | 7.32E-03 |
| $y = 7$ | 2.16E-02 |
| $y = 8$ | 5.39E-02 |
| $y = 9$ | 0.115 |
| $y = 10$ | 0.212 |
| $y = 11$ | 0.345 |
| $y = 12$ | 0.5 |
| ... | ... |

We chose to flip training and validation labels for samples whose predicted outcome was chosen correctly $y < 5$ out of N=25 times as the chance of occurrence is extraordinarily low at 0.0455% as shown in Supplement Table 2. Using this heuristic, 25% of GP3 and GP4 samples qualified as mislabeled using this Binomial criteria. The 25% of training/validation samples assumed to be mislabled were added to a "fliplist" that was later used to fine-tune the models.

Additionally, we decided to remove samples from the training/validation data sets that only classified correctly 5/25 to 9/25 times, as we deemed those samples to be ambiguous and only serve to confuse training. These samples were added to a ”blocklist” to eliminate these training and validation samples during subsequent fine-tuning.

Samples with better than 10/25 correct classifications were assumed to be probable events and preserved in the training/validation sets.

#### S3 - Confidence Calibration

Several techniques were tried to calibrate confidence scores, where the solution eventually employed convolving a Boolean array with a kernel window; the Parzen-Rozenblatt kernel window technique allowed for a smooth estimate of the confidence calibration function. Histogram binning is another popular approach, but the results seemed to exhibit much more epistemic noise between the various models. However, the expected calibration error (ECE) after histogram binning showed an improvement in the non-ensembled case to 8.6% and 8.3% in the ensembled case, which was slightly better than the Parzen-Rozenblatt kernel approach at 10.2% and 8.6%. Despite the lower ECE from histogram binning, the Parzen-Rozenblatt approach was chosen for the confidence remapping function since it produced a smoother translation and the results between models were more tightly clustered.

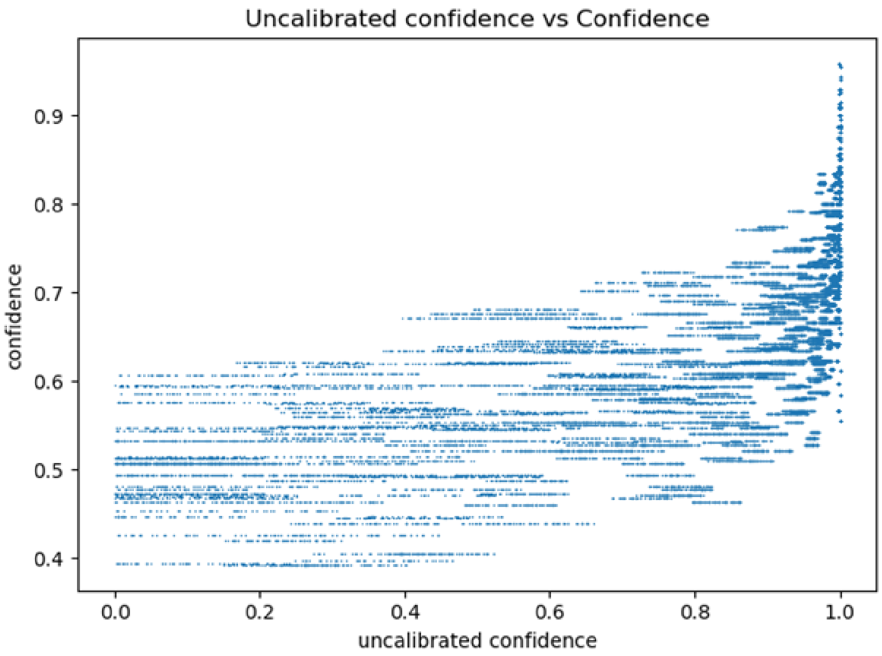


**Supplement Fig 1.** Histogram binning confidence calibration on non-ensemble output.

#### S4 - Confidence Filtering

We again measured performance on holdout for various *sample-fraction* thresholds from 0%-0.95%, which is shown in Supplement Table 3. Ignoring samples below the 35th percentile (indicated as 0.65 in Ignore column) produced our estimated improved accuracy=0.74, $F_1$=0.72 and AUC=0.79 performance shown in bold. If we had chosen to filter based on calibrated confidence $\hat{q}_i < 0.85$, our results would have been a couple of percentage points higher across the board as highlighted in gray (accuracy=0.76, $F_1$=0.74, and AUC=0.80); however, from the table we see that *sample-fraction* would have diminished to $< 0.2$, which we predicted from validation data is too aggressive and

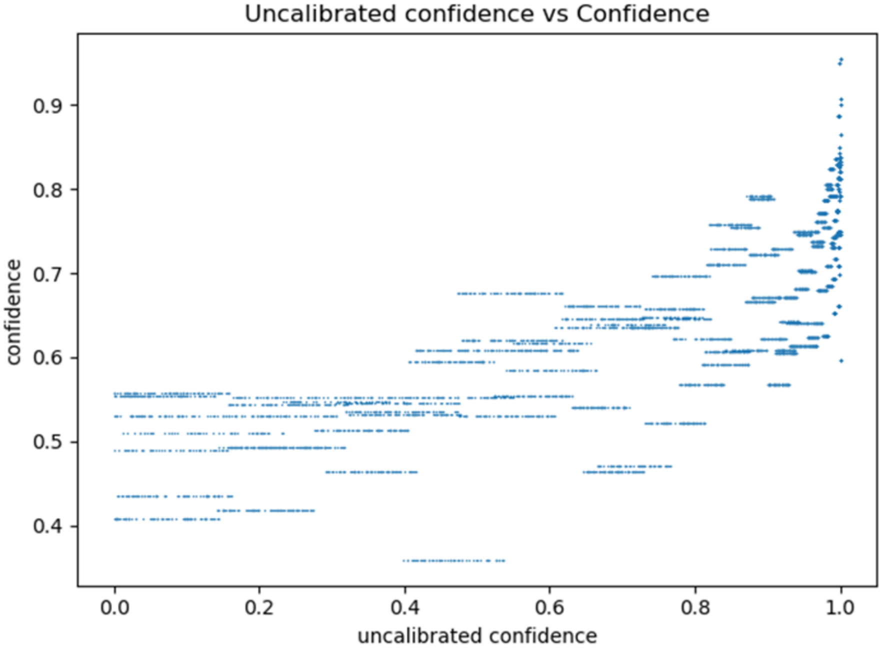


**Supplement Fig 2.** Histogram binning confidence calibration on ensemble output.

degrades performance. Tuning with larger data sets or other unseen sources can provide further evidence on which approach is better in practice. If the percentage of samples that map to a calibrated quality above the calibrated confidence threshold is very low, the sample may be considered out of distribution, and may be used to indicate careful consideration and inspection.

If we analyze the performance between the most and least confident samples for any threshold, we see a high degree of performance gain as shown in Supplement Fig 3 for the 65% threshold. This performance gain and significance is consistent across most thresholds except at very aggressive filtering (above 90%). The effect size (Cohen's d) and significance (Wilcoxon ranksum p-value) demonstrates that this approach is very effective for our pathology classification problem.

#### S5 - Total Optimization Effect

In section Results and sub-section Comparison to a Baseline in the main paper we show boxplots in Fig 8 comparing a "detuned" or unoptimized version of a VGG-16 and the optimized VGG-9 models; that comparison demonstrates un-ensembled detuned models to the tuned ensembled models. For completeness, here we also show two more comparisons for the unoptimized and optimized cases. First we show when both are un-ensembled in Supplement Fig 4, and when both are ensembled in Supplement Fig 5. It is clear with each of these comparisons that the unoptimized and optimized models show a very large performance gap.

#### S6 - Label Flipping on Holdout Test

The following discussion is speculative at best, and so none of the results shown below were provided in the formal paper. Instead the paper focuses on improving the results as best as possible while relying only on the original ground truth (GT) labels. There is some evidence that quite a few (25% GS3 and 29% GS4) of the epithelial glands in the

**Supplement Table 3.** VGG-9 holdout performance versus *sample-fraction*

| Ignored (Samp.Frac.=1-Ign.) | Accuracy | F1 | AUC | Confidence $\hat{Q}$ |
|---|---|---|---|---|
| 0.95 | 0.719 (0.12) | 0.704 (0.15) | 0.743 (0.12) | 0.873 (0.01) |
| 0.90 | 0.758 (0.08) | 0.745 (0.10) | 0.809 (0.06) | 0.866 (0.01) |
| 0.85 | 0.758 (0.09) | 0.737 (0.14) | 0.804 (0.07) | 0.856 (0.01) |
| 0.80 | 0.738 (0.10) | 0.706 (0.18) | 0.775 (0.10) | 0.831 (0.01) |
| 0.75 | 0.731 (0.10) | 0.700 (0.16) | 0.775 (0.09) | 0.828 (0.01) |
| 0.70 | 0.732 (0.09) | 0.699 (0.15) | 0.775 (0.09) | 0.821 (0.01) |
| **0.65** | **0.743 (0.08)** | **0.720 (0.12)** | **0.785 (0.08)** | **0.818 (0.01)** |
| 0.60 | 0.740 (0.07) | 0.712 (0.12) | 0.779 (0.07) | 0.805 (0.02) |
| 0.55 | 0.738 (0.08) | 0.707 (0.11) | 0.781 (0.07) | 0.798 (0.02) |
| 0.50 | 0.734 (0.07) | 0.705 (0.10) | 0.778 (0.06) | 0.785 (0.01) |
| 0.45 | 0.731 (0.07) | 0.704 (0.11) | 0.772 (0.07) | 0.779 (0.02) |
| 0.40 | 0.723 (0.07) | 0.697 (0.10) | 0.769 (0.06) | 0.758 (0.02) |
| 0.35 | 0.721 (0.06) | 0.698 (0.08) | 0.766 (0.05) | 0.743 (0.01) |
| 0.30 | 0.719 (0.06) | 0.697 (0.08) | 0.766 (0.05) | 0.733 (0.02) |
| 0.25 | 0.709 (0.06) | 0.688 (0.08) | 0.762 (0.04) | 0.713 (0.01) |
| 0.20 | 0.702 (0.06) | 0.680 (0.08) | 0.756 (0.05) | 0.695 (0.02) |
| 0.15 | 0.697 (0.06) | 0.675 (0.08) | 0.751 (0.05) | 0.667 (0.02) |
| 0.10 | 0.689 (0.05) | 0.668 (0.07) | 0.746 (0.04) | 0.642 (0.02) |
| 0.05 | 0.682 (0.05) | 0.660 (0.07) | 0.742 (0.04) | 0.610 (0.01) |
| 0.00 | 0.673 (0.05) | 0.651 (0.07) | 0.738 (0.04) | 0.524 (0.05) |

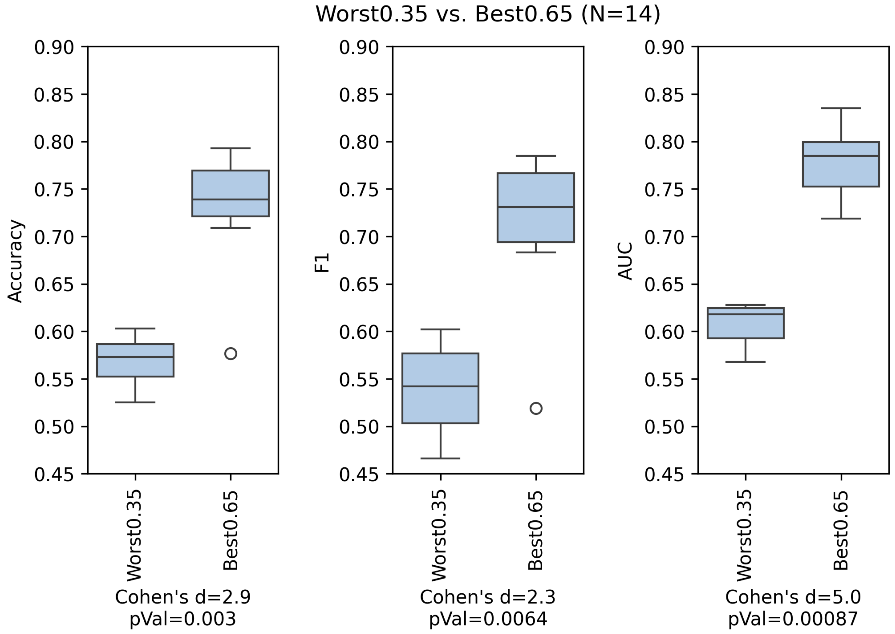


**Supplement Fig 3.** Most confident versus least confident performance at 65% threshold.

data cohort are better labeled contrary to GT. As shown in the Results Fig 6 of the formal paper, models were significantly improved in performance by training with refined labels. If we allot for flipped labels in the holdout data set, as expected we achieve a much higher performance. Importantly, this result is not based on the

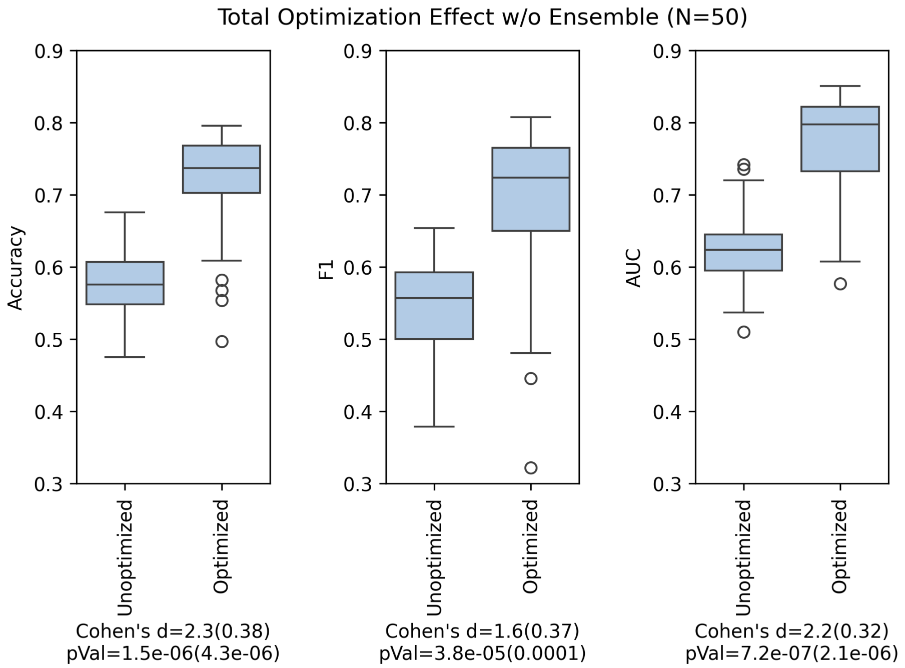


**Supplement Fig 4.** Non-ensemble comparison of baseline and optimized CNNs.

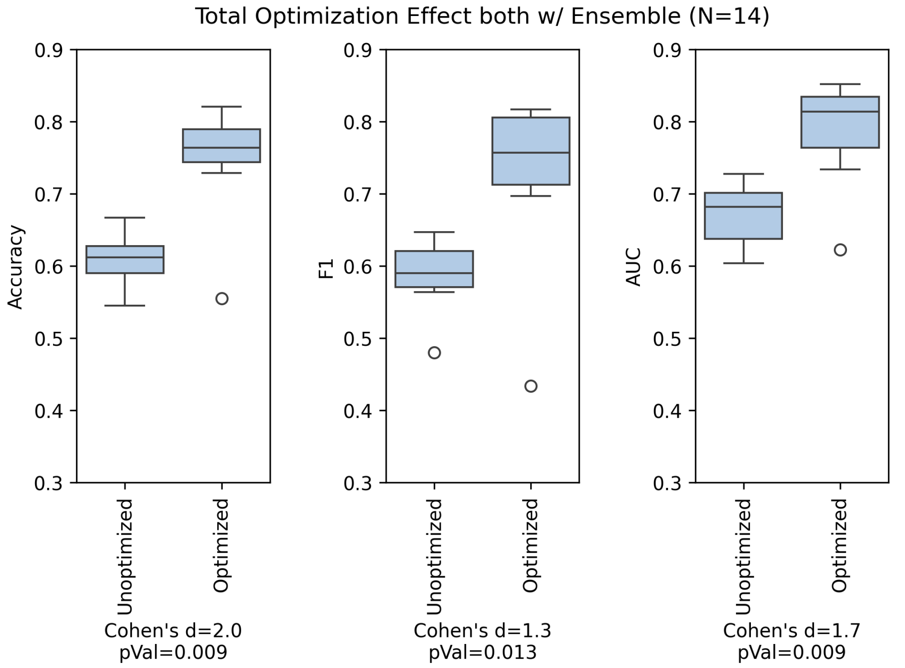


**Supplement Fig 5.** Ensemble Comparison of baseline and optimized CNNs.

inference decision of a model on the unseen holdout data (which would demonstrate perfect performance), but prior evidence that the holdout data label was in error and then tested on newly trained models.

The results in the tables below are collected by using all of the same optimization techniques aforementioned, including filtering 65% of the inferential samples with the lowest confidence, but additionally allowing for flipped labels based on prior evidence. The mean results averaged on all 35 models are shown in Supplement Table 4 and results from ensembles (ensembled across the 5x folds, and averaged across 7 shuffled tests) are shown in Supplement Table 5. Results are shown with slightly more precision here since AUC results are nearly 1.0. This result is optimistic, but we feel shows additional evidence that DL models can discriminate epithelial glands across Gleason 3 and 4 pattern areas with a high degree of discrimination.

**Supplement Table 4.** VGG9 performance on "refined" holdout set

| | Accuracy | F1 | AUC |
|---|---|---|---|
| Mean | **0.947** | **0.944** | **0.982** |
| Stddev | 0.039 | 0.044 | 0.018 |

**Supplement Table 5.** Ensembled VGG9 performance on "refined" holdout set

| | Accuracy | F1 | AUC |
|---|---|---|---|
| Mean | **0.971** | **0.970** | **0.993** |
| Stddev | 0.034 | 0.037 | 0.010 |
